# An Exploratory Study to Detect the Effects of the Combined Intake of Gamma-aminobutyric Acid (GABA) and L-theanine on Sleep by Wearable Device

**DOI:** 10.1101/2023.10.01.23296182

**Authors:** Hiroyuki Konno, Rikuto Murotani, Yukikazu Kamada

## Abstract

**Objectives:** Sleep disorders are a global issue, and supplements for sleep and new devices for daily sleep status assessment are becoming widely available. Gamma-aminobutyric acid (GABA) and L-theanine are commonly used dietary supplements to improve sleep. This study examined whether the combined GABA (700 mg/day) and L-theanine (200 mg/day) intake improve sleep in adults with sleep problems and whether the Fitbit Charge 5 can detect sleep status changes the supplements induced.

**Methods:** Participants received the supplements for four weeks, and changes in sleep quality measured using the Pittsburgh Sleep Quality Index (PSQI) and sleep-related data measured by the Fitbit Charge 5 were evaluated before and after the intake of the supplements.

**Results:** Results obtained from 19 participants indicated significant improvement in the total PSQI score (9.42 ± 1.80 to 6.26 ± 1.66 (mean ± standard deviation), p<0.001). Sleep score improvement was insignificant for the Fitbit data(N=17). However, sleep recovery scores improved significantly (p=0.042). In addition, heart rate during sleep decreased with a significant difference of 1.3 bpm decrease in the first week of intake (p = 0.045).

**Conclusions:** The simultaneous intake of GABA and L-theanine improved sleep in adults, and the Fitbit Charge 5 could detect improvements in objective information regarding sleep status.

## 1. Introduction

Sleep disorders are common worldwide and affect health and quality of life.^1^ Sleep disorders are among the most critical social problems in Japan. Epidemiological studies on insomnia in Japan have reported a prevalence of 14.6% in females and 12.2% in males.^2^ Epidemiological studies have reported the prevalence of difficulty initiating sleep, difficulty maintaining sleep with difficulty resuming sleep, and early morning awakening with difficulty resuming sleep as 11.0%, 8.1%, and 7.4%, respectively, in females and 8.3%, 5.8%, and 5.8%, respectively, in males. According to the epidemiological study of excessive daytime sleepiness in 30,000 subjects taken from the general population of Japan, difficulty initiating sleep, difficulty maintaining sleep, early morning awakening, and loss of deep sleep was 20.0%, 23.4%, 20.6%, and 24.8%, respectively, in females, and 14.2%, 18.1%, 26.9%, and 22.1%, respectively, in males.^3^ Mid-onset and early morning awakenings significantly increase in middle-aged and older adults.^4^ Lack of sleep and poor sleep quality significantly impact health negatively and cause significant stress, which in the long term may be a risk factor for lifestyle-related diseases and psychiatric disorders such as depression and dementia.^5-7^ Furthermore, poor sleep and sleep disorders are known to reduce performance and productivity at work.^8,9^ A cross-sectional study in Japan has reported that sleep health is associated with presenteeism.^10^ Sleep drugs are mainly administered at medical institutions to treat insomnia. However, long-term administration can lead to the development of dependence, making it difficult to stop taking medication.^11^

Gamma-aminobutyric acid (GABA) is an amino acid found in many foods. GABA is a neurotransmitter with anxiolytic and hypnotic effects,^12^ which is expected to improve sleep and stress.^13^ Clinical trials have reported that in Japanese adults with sleep disorders, 100 mg of GABA reduced the time required to fall asleep, prolonged non-REM sleep time, and improved mood during waking.^14^ L-theanine is an amino acid unique to green tea and has been reported to have anti-anxiety and sleep-improving effects in clinical studies.^15^ GABA and L-theanine are widely used commercially as dietary supplements to improve sleep quality, with few problems related to dependence or long-term continuous use.^16,17^ However, there is a report in mice on the efficacy of GABA and L-theanine on sleep when taken simultaneously,^18^ which was higher when combined than when each was administered alone. There is no clinical evidence for the efficacy and safety of the combination.

In addition, wearable device sleep assessment technology has been developed and put to practical use in recent years. Among these, Fitbit has been validated and used for sleep assessments in many clinical trials.^19^

In this study, an exploratory single-arm before-and-after study was conducted to evaluate the effects of GABA and L-theanine for four weeks on sleep and assess whether a wearable device could detect the effects of GABA and L-theanine intake.

## 2. Materials and Methods

This study was conducted according to the Declaration of Helsinki and approved by the Institutional Review Board of Brain Care Clinic (BCC230203-2, date of approval: February 3, 2023, UMIN Study ID: UMIN000050613). This study was conducted between February and April 2023 and included Japanese adults with sleep-related problems.

### Participants

The inclusion and exclusion criteria were as follows:

Inclusion criteria

1. Males and females aged 20–64 were aware of sleep-related problems.
2. BMI between 18.5 and 25.0 kg/m2.
3. Those with a score of 6 or more on the Japanese version of the Pittsburgh Sleep Quality Index(PSQI-J).
4. Being able to wear a wearable device (Fitbit Charge 5) during the study period, including at bedtime.
5. Voluntary participation

Exclusion criteria.

1. Patients currently diagnosed with insomnia or sleep disorders at a medical institution undergoing treatment.
2. Consumption of foods or health foods containing GABA or L-theanine.
3. Regular consumption of healthy foods or sleep supplements
4. Serious diseases such as glucose metabolism-related, lipid metabolism-related, liver, renal, cardiovascular, gastrointestinal, respiratory, hematological, autoimmune, endocrine, and metabolic diseases. Alternatively, those with a history of such diseases may be excluded.
5. History of drug or severe food allergies.
6. Those with abnormal laboratory test results within the past year were considered to have problems participating in the trial.
7. Those with a disease underwent hospital visits, treatment, medication use, or follow-up.
8. Those judged to be unsuitable based on the results of the screening survey.
9. Those participating in other clinical studies at the start of the study.
10. Those who are pregnant or plan to become pregnant or breastfeed during the study period.
11. Those who are deemed unsuitable for participation by the study investigator.

### Study procedure

Investigators recruited adults who met the inclusion criteria via a web-based questionnaire and mailed them a consent document and screening questionnaire. Participants received an explanation from the investigators via a web interview or telephone call, signed a consent document, completed a screening questionnaire four weeks before supplement intake, and mailed the consent document and screening questionnaire to the investigators. The screening survey included the PSQI-J, daily activities, and most recent physical condition. Participants who qualified for screening visited the study site two weeks before the start of intake to perform the State-Trait Anxiety Inventory (STAI), measure their weight, and receive supplements (GABA and L-theanine), a Fitbit Charge 5, a smartphone for Fitbit Charge 5 synchronization, and a daily diary. The Fitbit Charge 5 was worn for two weeks before the start of intake, and participants were instructed to wear the Fitbit Charge 5 daily, except when bathing. A daily diary was kept to record changes in physical condition, daily activities, such as alcohol consumption and exercise, medication intake, and test supplements (GABA and L-theanine) taken from the start of supplement intake. The participants took the study supplements GABA 700 mg and L-theanine 200 mg (PURE Encapsulations, Nestle Health Science, Tokyo, Japan) once daily before bedtime with 150 mL of water. Participants visited the study site four weeks after supplementation and underwent the PSQI-J, STAI, and weight measurements.

### Measurements

The PSQI-J was used as the primary endpoint in this study.^20-22^ The cutoff value for the total PSQI (PSQI-J global) was 5.5 points, and a score of 6 or more was defined as having a sleep problem. The secondary endpoints included STAI and personal health record data from the Fitbit Charge 5, including sleep duration, sleep score, heart rate during sleep, step count, and skin electrical stimulation activity. According to the Fitbit Charge 5, sleep time was calculated by subtracting the time spent awake or restless sleep from the total time recorded.^23^ In addition to the total sleep time, sleep time is classified into three stages: REM sleep time, deep sleep time, and light sleep time. Sleep scores were calculated on a 100-point scale according to sleep duration, sleep stage, and sleep recovery (how relaxed an individual is during sleep). The sleep recovery score was calculated on a 25-point scale according to the heart rate during sleep, tossing, and turning. The heart rate during sleep was detected continuously throughout the night and was calculated as the average value (bpm) over one night.^23^ Electro Dermal Activity, a stress-related indicator, is calculated by sensing changes in the electrical activity of skin perspiration, and the Fitbit Charge 5 detects the skin’s electrical activity response at the fingertips.^23^ Measurements were performed once daily. Adverse events were included as secondary endpoints. The principal investigator assessed the causal relationship between the adverse events and study supplementation.

### Data Analysis

Statistical analysis was performed using R version 4.2.0.; PSQI, STAI, and Fitbit Charge 5 data were compared before and after the start of the study supplement intake, and p-values were calculated using a paired t-test, assuming normality and equal variance in the data. A comparison of sleep-related data from the Fitbit Charge 5 before and after the analysis was performed in a population (Group B), whereas cases with a high number of missing data (three missing days in a week, i.e., missing sleep scores of more than 40%) were excluded, compared with the mean values for the seven days before and after intake of the study supplements. The significance level was set at 5%. Daily sleep time and step count data obtained on a Fitbit Charge 5 are shown as line charts for each case, and scatter plots of sleep time and step count data were also created.

## 3. Results

### Participant Characteristics

Twenty participants were eligible for the supplement intake intervention in this study, and all 20 completed the study, with one participant’s intake compliance falling below 85% (23/28 days, 82.1%) (Figure 1). The subjects’ background information is shown in Table 1.

**Figure 1.**
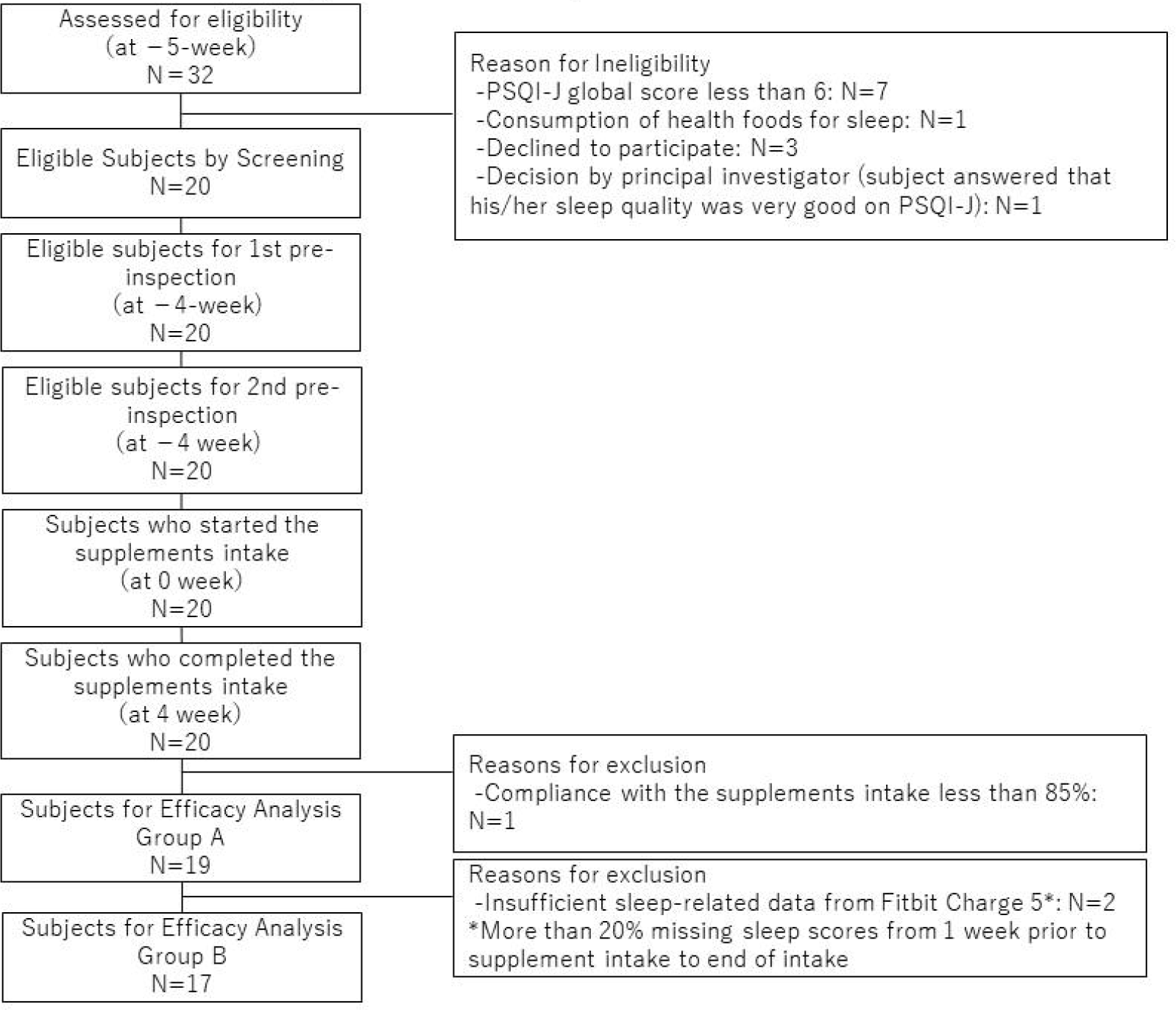
The study flowchart.

**Table 1.** Participants characteristics.

| Variables | Total<br>(N=20) |
| --- | --- |
| Age (years) | 48.4 (24 - 63) |
| Gender | Female 10: Male 10 |
| Work status | Working 19: Not-working 1 |
| Desk worker | 12 |
| Physical labor worker | 6 |
| Manual laborer | 1 |
| Body mass index (kg/m<sup>2</sup>) | 22.2 (18.9 – 24.9) |
| PSQI-J global before intake of the supplement | 9.25 (6 - 12) |

N=20
Mean (Min-Max)

### Efficacy Evaluation

The PSQI-J global was 9.42 ± 1.80 (mean ± standard deviation) before intake of the study supplement. It significantly improved to 6.26 ± 1.66 after four weeks of continued intake (p<0.001) (Figure 2, Table S1). In addition, before intake of the study supplement, all 19 patients had a score of 7 or above, and 10 had a PSQI-J global score of 10 or above; 16 of the 19 patients (84.2%) showed an improvement of 1 or more points. After intake, six patients (31.6%) had a score of 5 or below, considered without sleep problems, and six patients (31.6%) had a score of 6. Furthermore, the PSQI-J global score in Group B improved significantly from 9.29 ± 1.86 before intake to 6.18 ± 1.70 after intake (p<0.001). Significant improvements were observed in Subjective sleep quality (C1), Sleep latency (C2), Sleep duration (C3), and Daytime dysfunction (C7) (Table 2, Figure S1).

**Table 2.** The Pittsburgh sleep quality index.

| Items | Before | After | P-value |
| --- | --- | --- | --- |
| Component 1:<br>Subjective sleep quality | 2.16 ± 0.37 | 1.42 ± 0.51 | <0.001* |
| Component 2:<br>Sleep latency | 2.32 ± 0.75 | 1.79 ± 0.85 | 0.002* |
| Component 3:<br>Sleep duration | 1.89 ± 0.57 | 1.37 ± 0.83 | 0.014* |
| Component 4:<br>Habitual sleep efficiency | 0.84 ± 0.96 | 0.53 ± 0.61 | 0.055 |
| Component 5:<br>Sleep disturbances | 0.95 ± 0.23 | 1.00 ± 0.00 | 0.331 |
| Component 6:<br>Use of sleeping medicine | 0.00 ± 0.00 | 0.00 ± 0.00 | NaN |
| Component 7:<br>Daytime dysfunction | 1.26 ± 0.56 | 0.16 ± 0.37 | <0.001* |
| PSQI-J global (0-21) | 9.42 ± 1.80 | 6.26 ± 1.66 | <0.001* |

N=19
NaN: Not a Number , PSQI-J: Japanese version of the Pittsburgh Sleep Quality Index
Mean ± Standard Deviation
P-values were calculated using a paired t-test, with a significance level of 5%

**Figure 2.**
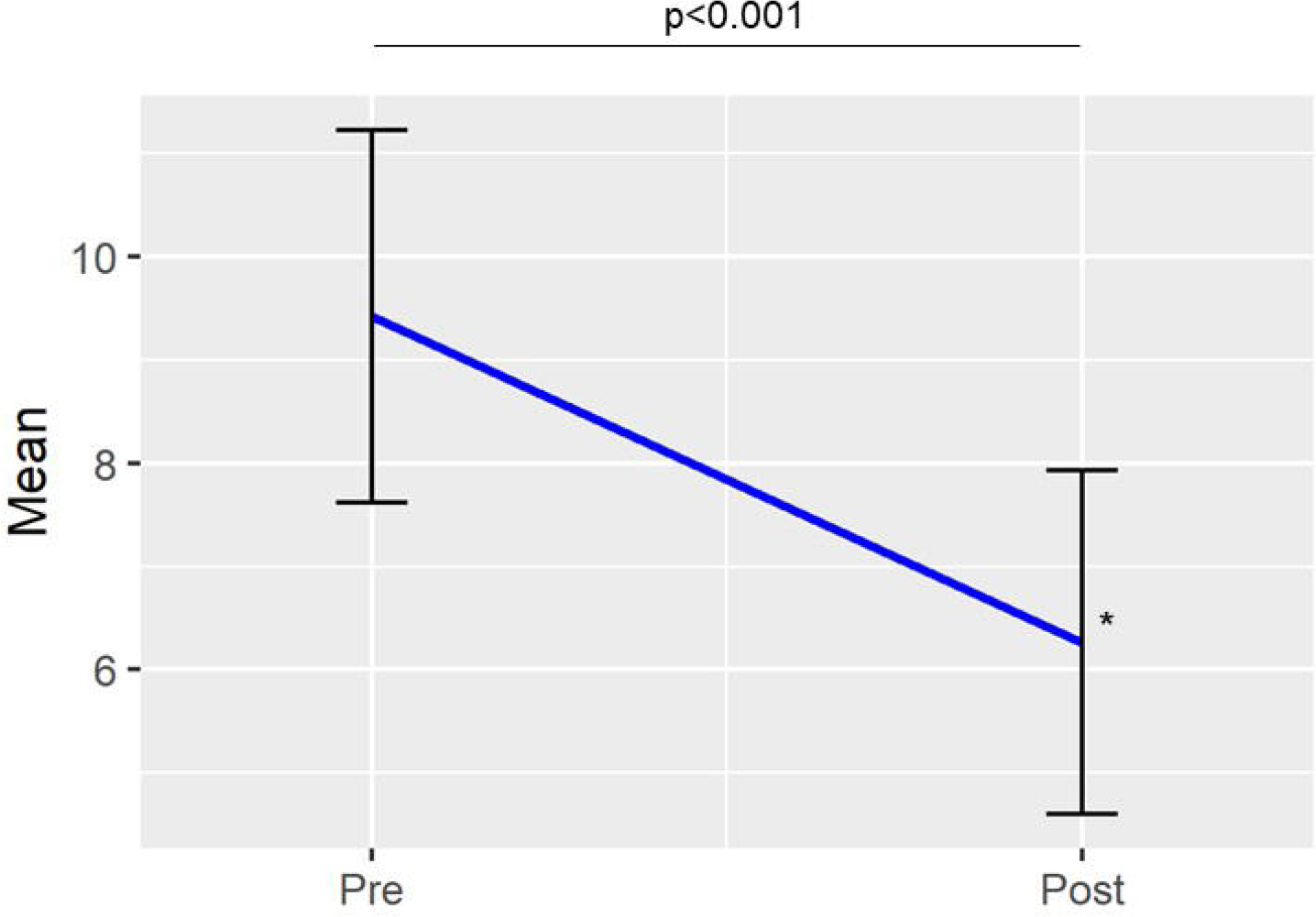
A Japanese version of the Pittsburgh Sleep Quality Index global score N=19 Mean ± standard deviation. P-values were calculated with a paired t-test: level of significance 5%.

The STAI scores were lower after intake than before study supplementation but were not significantly different (Table 3).

**Table 3.**
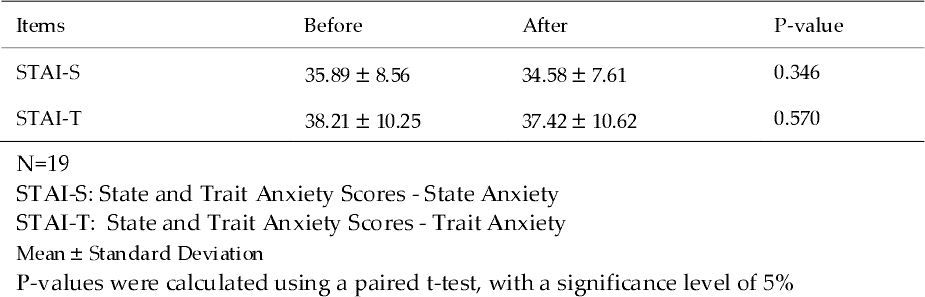
State and Trait Anxiety Scores.

Statistical comparisons were not performed before and after supplement intake because of the large amount of missing data on sleep duration and sleep stage collected by Fitbit Charge 5, but significant changes were not observed before and after supplement intake (Figure S2). No correlation was found between the number of steps taken and the sleep duration or stage (Figure S3). In Group B, sleep scores were better after the intake of the study supplement than before, but there were no statistical differences. The sleep recovery score showed a significant improvement of 0.76 points after four weeks of intake compared to the pre-test supplementation (p=0.042) (Figure 3). The heart rate during sleep decreased at each post-test week, with a significant difference of 1.3 bpm decrease in the first week of intake (p = 0.045) (Figure 3). The electrodermal activity, a stress-related measure, decreased in each post-test week compared to before, with a significant 1.0 decrease in the first week of supplementation (p=0.029) (Figure 3).

**Figure 3.**
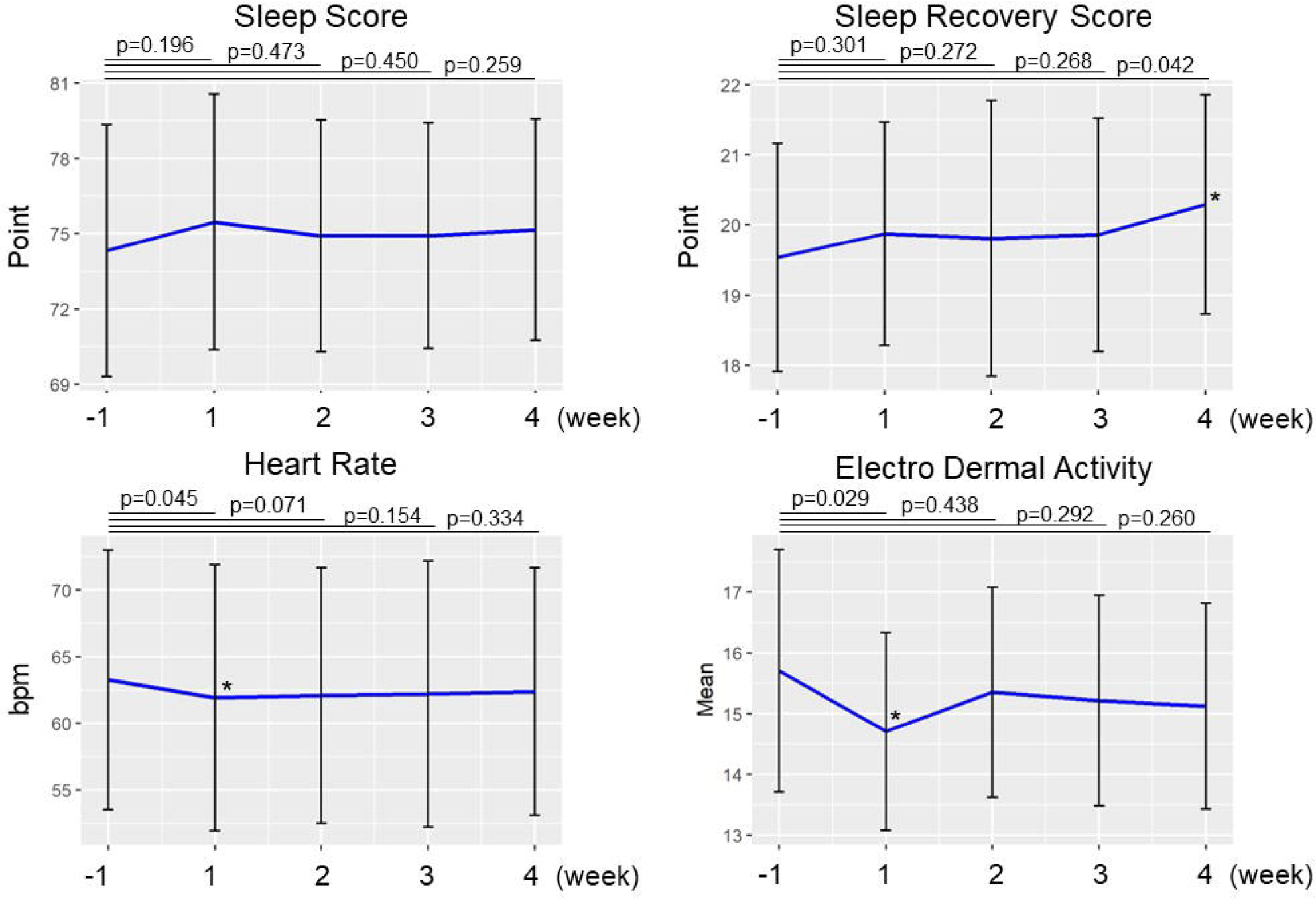
Sleep-related data measured by Fitbit Charge 5. N=17 Mean ± standard deviation. P-values were calculated with a paired t-test: level of significance 5%.

### Adverse Events

Adverse events occurred in four individuals during the study period. There were three cases of headache, one case of heaviness of the head, one case of diarrhea, and one case of thorn puncture wound, all of which lasted less than one day, except for three days of diarrhea. The principal investigator judged that there was no causal relationship between the study supplement and the occurrence of adverse events, as all adverse events resolved during continued GABA and L-theanine intake.

## 4. Discussion

This study showed that simultaneous intake of GABA and L-theanine improved sleep problems in adults and that the Fitbit Charge 5 could detect and evaluate objective improvements in sleep status. GABA and L-theanine have been reported to improve sleep.^13-15^ The GABA 700 mg/capsule used in this study is the highest dose of GABA supplement available in Japan, and its intake in combination with L-theanine significantly improved the global PSQI-J score by more than three points. According to reports from previous clinical trials in Japanese participants with sleep-related problems without a diagnosis of insomnia, GABA at 100 mg/day for one week improved the PSQI by less than one point,^14^ and L-theanine at 200 mg/day for four weeks improved the PSQI by 1.37 point.^15^ Although it is difficult to interpret the results because of the difference in design and GABA dose in this study, the improvement in PSQI in this study was larger than that in previous reports, demonstrating the possibility of a synergistic or additive effect of the combination of GABA and L-theanine. This study also recommends that GABA (700 mg/day) and L-theanine (200 mg/day) could be administered concomitantly without any safety issues.

The most significant impact of this study was that the PSQI, one of the most used sleep assessment tools in research and clinical practice in the field of sleep, and the Fitbit Charge 5 were used simultaneously to assess sleep, and the effectiveness of the combined intake of GABA and L-theanine on sleep was detected in both assessments. Several clinical trials have been conducted on sleep using Fitbit.^24-26^ However, there are no reports of clinical trials in which Fitbit was used to evaluate the effects of nonpharmaceutical supplements on sleep. In this study, the combination of GABA and L-theanine significantly improved the PSQI and Fitbit Charge 5 sleep recovery scores. The sleep score showed no statistically significant difference, although numerical increases were observed. Sleep duration and stage factors were included in the sleep score calculation, and the result of the sleep score may have been affected by many missing measurements of these factors. The sleep recovery score was calculated using the heart rate, tossing, and turning. This score could also be associated with a significant improvement in PSQI Component 1, subjective sleep quality. The sleep heart rate significantly decreased in the first week after intake and remained stable at lower values than before intake in the second week and after that. Heart rate, which reflects the autonomic nervous system, is considered to be related to the sleep stage,^27^ and the combined intake of GABA and L-theanine may have influenced the improvement of sleep quality.

GABA and L-theanine have been reported to be effective against anxiety and stress,^13, 15^ but STAI did not significantly improve in this study. This lack of improvement is thought to be because anxiety was not a selection criterion. The STAI scores before taking the study supplement were not as high as 35.89 for state anxiety and 38.21 for trait anxiety; therefore, no improvement was detected. In contrast, electrodermal activity measured with a Fitbit Charge 5 showed significant improvement after one week. Electrodermal activity is a method of electrically measuring sympathetic sweat gland activity. It indicates the autonomic nervous system,^28^ showing that GABA and L-theanine affect daily stress, as detected by the Fitbit Charge 5.

In this study, no statistical analysis was performed on the sleep duration and duration of each sleep stage based on the Fitbit Charge 5 because there were many missing measurements, and the study was limited to observing trends on a case-by-case basis. According to the results, there were no major changes in sleep duration based on the Fitbit Charge 5 detection or the duration of each sleep stage before and after the combined intake of GABA and L-theanine. In addition, the number of steps taken was employed as daily activity data and analyzed for correlation with sleep duration. However, no correlation was observed, like a report by Guillaum et al.,^29^ suggesting that the intensity of daytime activity did not affect sleep-related outcomes in this study.

The limitation of this study is that it was an exploratory study with a single arm before and after comparison. The primary endpoint was a questionnaire based on subjective judgments, and the number of participants was limited. It is necessary to investigate the efficacy of GABA and L-theanine on sleep by conducting a randomized controlled trial with a larger number of participants and using the Fitbit Charge 5, which can objectively evaluate sleep conditions without missing any sleep-related data.

## Data Availability

All relevant data are presented in this manuscript. All the data are available from the authors upon request.

## Supplementary figures

Figure S1. The seven components of the Pittsburgh sleep quality index

N=19

P-values were calculated using a paired t-test, with a significance level of 5%.

The graph for Component 6: Use of sleeping medicine was not drawn because all patients had a score of 0 before and after according to the exclusion criteria of the study.

Figure S2. Daily sleep time and step count data obtained on a Fitbit Charge 5 Figure S3. Scatter plots of number of steps and sleep time

N=20

Left: Daily values for 1 week before supplementation

Right: Daily values for 4 weeks after supplementation

Horizontal axis: number of steps

Vertical axis: sleep time (minutes)

## Acknowledgments

We thank Ms. Chie Matsuda (Google Japan) for helping us with information on the Fitbit Charge 5.

## Informed Consent Statement

Informed consent was obtained from all participants involved in the study.

## Registry and the Registration No. of the study

UMIN000050613

