## Supplementary figures and images for "An Exploratory Study to Detect the Effects of the Combined Intake of Gamma-aminobutyric Acid (GABA) and L-theanine on Sleep by Wearable Device"

### Figure S1

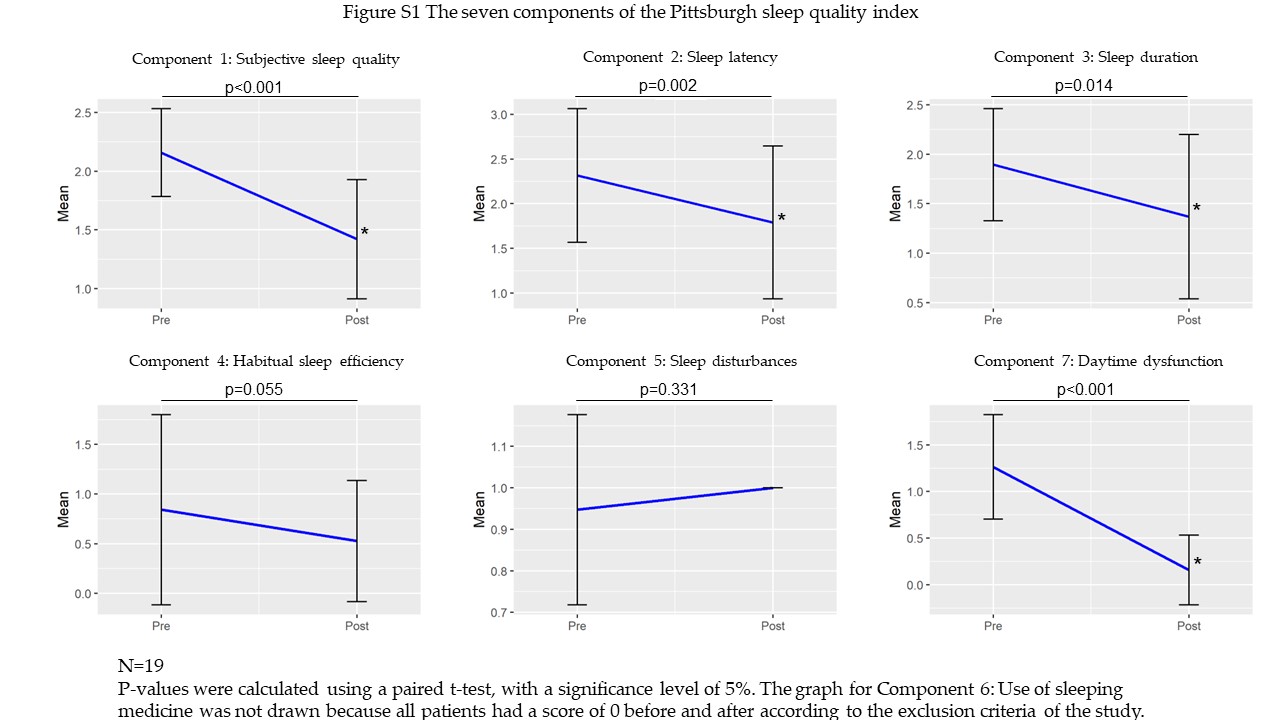

### Figure S2

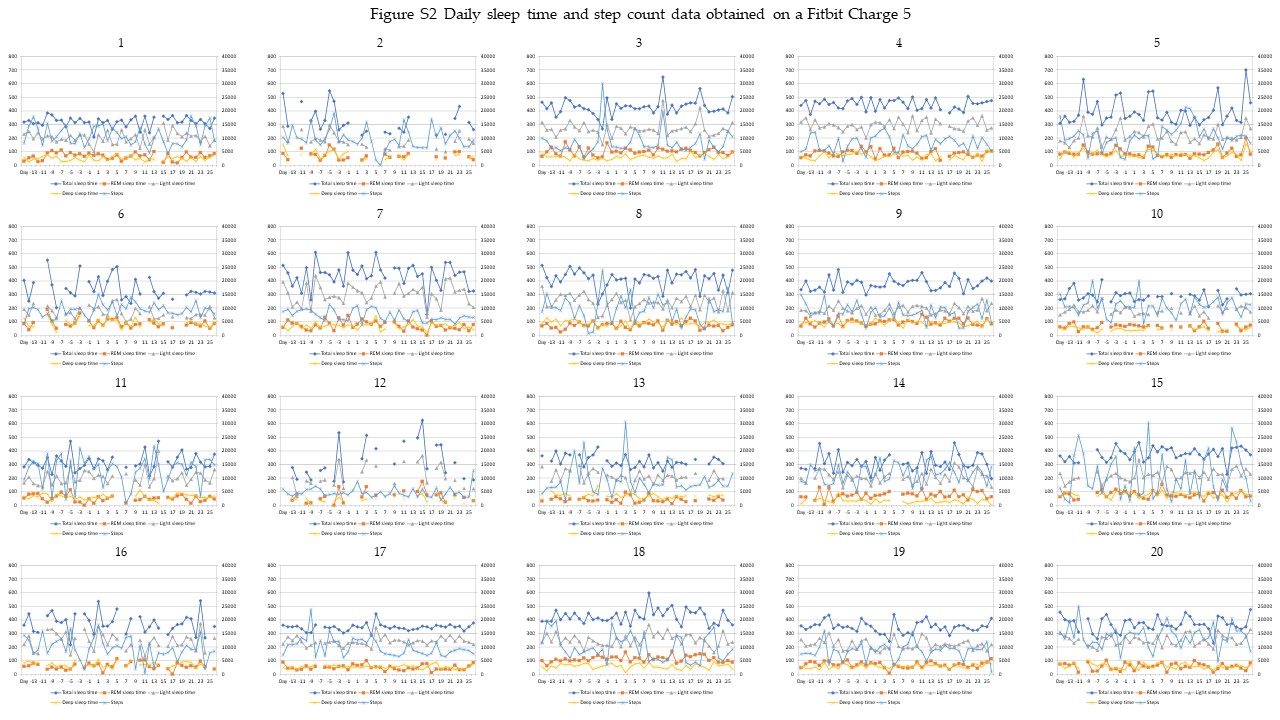

### Figure S3

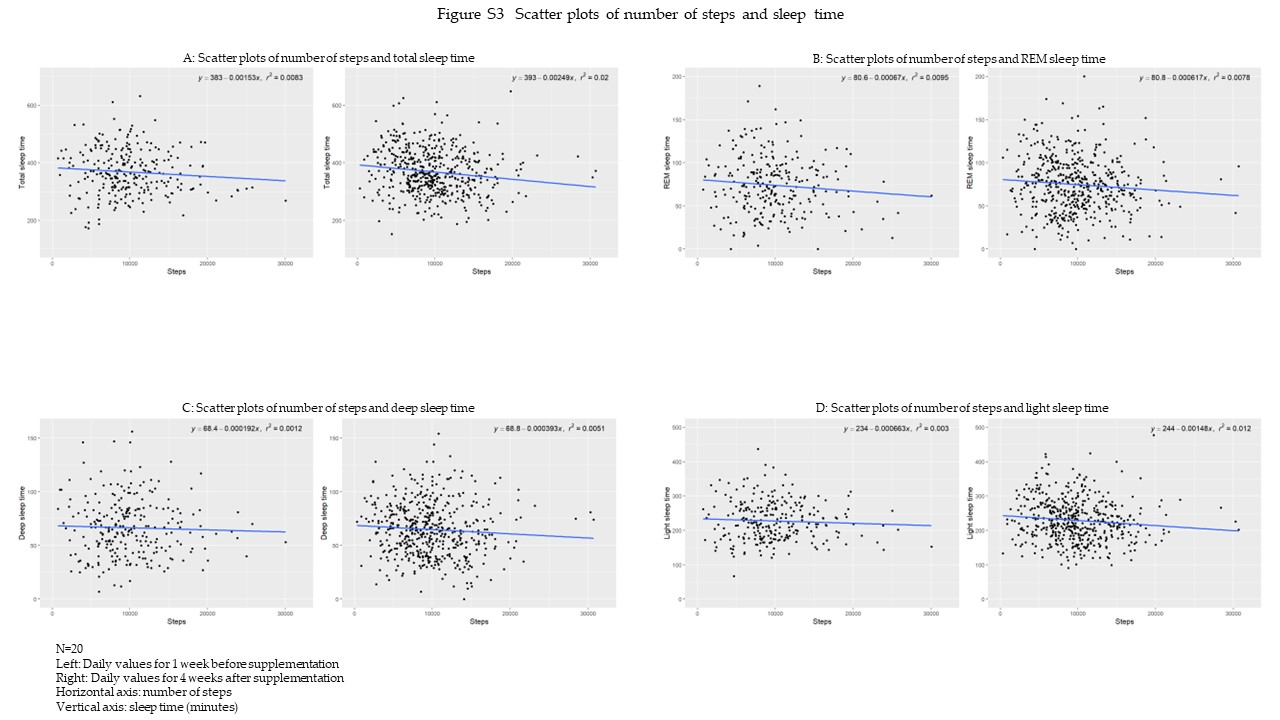
